# Dramatic impact of rapid point of care nucleic acid testing for SARS-CoV-2 in hospitalised patients: a clinical validation trial and implementation study

**DOI:** 10.1101/2020.05.31.20114520

**Authors:** Dami Collier, Sonny M. Assennato, Ben Warne, Nyarie Sithole, Katherine Sharrocks, Allyson Ritchie, Pooja Ravji, Matthew Routledge, Dominic Sparkes, Jordan Skittrall, Anna Smielewska, Isobel Ramsey, Neha Goel, Martin Curran, David Enoch, Rhys Tassell, Michelle Lineham, Devan Vaghela, Clare Leong, Hoi Ping Mok, John Bradley, Kenneth GC Smith, Vivienne Mendoza, Nikos Demiris, Martin Besser, Gordon Dougan, Paul J. Lehner, Mark J. Siedner, Hongyi Zhang, Claire S. Waddington, Helen Lee, Ravindra K. Gupta, and the CITIID-NIHR COVID BioResource Collaboration

## Abstract

**Background:** There is urgent need for safe and efficient triage protocols for hospitalized COVID-19 suspects to appropriate isolation wards. A major barrier to timely discharge of patients from the emergency room and hospital is the turnaround time for many SARS-CoV-2 nucleic acid tests. We validated a point of care nucleic acid amplification based platform SAMBA II for diagnosis of COVID-19 and performed an implementation study to assess its impact on patient disposition at a major academic hospital.

**Methods:** We prospectively recruited COVID-19 suspects admitted to hospital (NCT04326387). In an initial pilot phase, individuals were tested using a nasal/throat swab with the SAMBA II SARS-CoV-2 rapid diagnostic platform in parallel with a combined nasal/throat swab for standard central laboratory RT-PCR testing. In the second implementation phase, we examined the utility of adding the SAMBA platform to routine care. In the pilot phase, we measured concordance and assay validity using the central laboratory as the reference standard and assessed assay turnaround time. In the implementation phase, we assessed 1) time to definitive bed placement from admission, 2) time spent on COVID-19 holding wards, 3) proportion of patients in isolation versus COVID negative areas following a test, comparing the implementation phase with the 10 days prior to implementation.

**Results:** In phase I, 149 participants were included in the pilot. By central laboratory RT-PCR testing, 32 (21.5%) tested positive and 117 (78.5%). Sensitivity and specificity of the SAMBA assay compared to RT-PCR lab test were 96.9% (95% CI 0.838-0.999) and 99.1% (0.953-0.999), respectively. Median time to result was 2.6 hours (IQR 2.3 to 4.8) for SAMBA II SARS-CoV-2 test and 26.4 hours (IQR 21.4 to 31.4) for the standard lab RT-PCR test (p<0.001). In the first 10 days of the SAMBA implementation phase, we conducted 992 tests, with the majority (59.8%) used for hospital admission, and the remainder for pre-operative screening (11.3%), discharge planning (10%), in-hospital screening of new symptoms (9.7%). Comparing the pre-implementation (n=599) with the implementation phase, median time to definitive bed placement from admission was reduced from 23.4 hours (8.6-41.9) to 17.1 hours (9.0-28.8), *P*=0.02 in Cox analysis, adjusted for age, sex, comorbidities and clinical severity at presentation. Mean length of stay on a COVID-19 ‘holding’ ward decreased from 58.5 hours to 29.9 hours (*P*<0.001). Use of single occupancy rooms amongst those tested fell from 30.8% before to 21.2% (*P*=0.03) and 11 hospital bay closures (on average 6 beds each) were avoided after implementation of the POC assay.

**Conclusions:** The SAMBA II SARS-CoV-2 rapid assay performed well compared to a centralized laboratory RT-PCR platform and demonstrated shorter time to result both in trial and real-world settings. It was also associated with faster time to definitive bed placement from the emergency room, greater availability of isolation rooms, avoidance of hospital bay closures, and greater movement of patients to COVID negative open “green” category wards. Rapid testing in hospitals has the potential to transform ability to deal with the COVID-19 epidemic.

## Introduction

As of June 1^st^ 2020 there were over 400, 000 deaths worldwide and 40,000 deaths in the UK attributed to COVID-19^1^. Current clinical testing for acute SARS-CoV-2 infection and infection risk relies on nucleic acid detection using reverse transcription polymerase chain reaction (RT-PCR) on nose/throat swabs^2,3^. Antibodies to SARS-CoV-2 are detectable in only approximately 50% of infected people by day 5-7 after symptom onset,^4^ and are therefore not suitable as a test for early stages of infection, although they may add value in the second phase of illness when virus detection wanes in upper respiratory tract samples^3,5^. Nucleic acid testing usually requires central laboratory testing with concomitant delays, and turnaround times are usually in excess of 24 hours, and often days^6^.

Due to diverse presentations of COVID-19^7^, lack of a timely diagnosis can have serious consequences, including deadly nosocomial outbreaks^8,9^. Moreover, identifying and isolating patients appropriately as suspects or cases is critical for hospital flow and resource allocation. Misclassifying cases as non-cases puts patients and healthcare providers at risk. Conversely, misclassifying patients without disease as suspects increases use of scarce personal protective equipment and isolation wards. Therefore, screening hospital admissions rapidly is critical to manage patient flow and limit potential for nosocomial transmission^10,11^. In the absence of a reliable rapid assay, hospitals have resorted to creating bespoke care pathways to use isolation rooms most effectively for COVID-19 suspects without a confirmed diagnosis^12^. Finally, given care home outbreaks, there is also urgent need to rapidly demonstrate COVID-19 status on discharge planning^13^. This need for rapid and safe patient movement is likely to increase sharply in late 2020 when norovirus and influenza (with or without SARS-CoV-2^14^) will likely compound pressure on hospitals and isolation capacity in particular. Such an approach would also relieve pressure on hospital virology laboratories so they can resume routine testing.

One potential solution to facilitating rapid patient triage and room allocation is to improve efficiency of COVID-19 testing. A number of near patient tests for SARS-CoV-2 have been described. Some have not performed well^15^, and none have undergone testing under rigorous clinical trial conditions with ‘real world’ data on impact on patient managment^16–20^.

Thorough, prospective evaluation for a high consequence pathogen such as SARS-CoV-2 is particularly important given risks related to false positives or negatives in the hospital setting. SAMBA (simple amplification based assay), an isothermal amplification based platform, has been extensively field tested for HIV diagnostic applications in low resource settings^21,22^, and has been adapted for use in SARS-CoV-2 with successful pre-clinical testing using synthetic standards and stored positive and negative clinical samples^23^. Here we present a prospective clinical validation study comparing SAMBA II SARS-CoV-2 performance against the standard laboratory RT-PCR test in suspected COVID-19 cases presenting to hospital, followed by analysis of POC implementation in hospital.

## Methods

### Assay validation study

The COVIDx Study was a prospective, comparative, real world assessment of SAMBA II SARS-CoV-2 point of care testing compared to the standard laboratory RT-PCR testing in participants admitted to Cambridge University Hospitals NHS Foundation Trust (CUH) with a possible diagnosis of COVID-19 (NCT04326387). CUH is a 1200-bed hospital providing secondary care to a population of 580,000 people in Cambridge and the surrounding area, as well as tertiary referral services to the East of England.

#### Validation Phase Participants

For the Phase I assay validation period, recruitment started two weeks into the national lockdown implemented by the UK government in response to the pandemic. Eligible consecutive participants were recruited during 12-hour day shifts over a duration of 4 weeks from the 6^th^ of April 2020 to the 2^nd^ of May 2020. We recruited adults (>16 years old) presenting to the emergency department or acute medical assessment unit and designated as COVID-19 suspects. This included participants who met the Public Heath England (PHE) definition of a possible COVID-19 case (see supplemental methods). The inclusion criteria were later expanded to include any adult requiring hospital admission and who was symptomatic of SARS-CoV-2 infection, demonstrated by clinical or radiological findings. This was done due to the changing landscape of the COVID-19 epidemic and emergence of new symptoms such as anosmia and diarrhoea. Exclusion criteria included not having the standard lab RT-PCR test applied within an 18-hour window of SAMBA SARS-CoV-2 test and those unwilling or unable to comply with study swabbing procedures.

#### Assay methods

Participants in the assay validation phase underwent testing with SAMBA II SARS-CoV-2 using a combined nasal/throat swab within 18 hours of a similar assay for the standard laboratory RT-PCR test. The index test is the SAMBA II SARS-CoV-2 Test, a nucleic acid amplification test (NAAT) which uses nucleic acid sequence based amplification to detect SARS-CoV-2 RNA from throat and nose swab specimens collected by dry sterile swab and inactivated in a proprietary inactivation buffer prior to analyses. This obviates the need for a BSL3 laboratory for specimen handling or viral extraction. The SAMBA II SARS-CoV-2 targets 2 genes-Orf1 and the E genes. The limit of detection (LoD) of the SAMBA II SARS-CoV-2 Test is published as 250 copies/mL^23^. The reference test is an in-house RT-PCR test developed in the public health England (PHE) laboratory at CUH with similar LOD.

Indeterminate SAMBA II SARS-CoV-2 tests were repeated with a 1:2 dilution of sample to inactivation buffer according to manufacturer standard operating procedures until a valid result was obtained. Indeterminate standard lab RT-PCR tests were repeated on a replicate nose/throat swab until a valid result was obtained. The results of the SAMBA II SARS-CoV-2 was not known to the assessors of the standard lab RT-PCR prior.

Demographic and clinical data were obtained at presentation from the hospital’s electronic patient records (EPIC) and entered into anonymised case report forms on the MACRO electronic database. Biological specimens from a combined nose and throat swab were collected and stored by research nurses. Results of the SAMBA assay were not made available to clinical teams during the pilot validation study. The primary outcome measures were time to result, concordance with the standard lab RT-PCR test and sensitivity/specificity of the SAMBA II SARS-CoV-2 test compared to the central laboratory RT-PCR assay.

#### Validation Study Sample Size and Analysis

We assumed a target sensitivity of 0.95 and disease prevalence of 15%. Using a 5% significance level and allowing for a precision of +/-5% gave a required sample size of 122. Participants with missing SAMBA II SARS-CoV-2 or standard lab RT-PCR tests result were excluded from the analyses. Descriptive analyses of clinical and demographic data are presented as median and interquartile range (IQR) when continuous and as frequency and proportion (%) when categorical. The difference in continuous and categorical data were tested using Wilcoxon rank sum and Chi-square test respectively. Agreement between the two tests was assessed using Cohen’s kappa, a correlation-like measure which accounts for agreement by chance alone, in which case κ = 0, while κ = 1 and κ = −1 correspond to perfect agreement and completely discordant pairs respectively. Sensitivity and specificity of SAMBA II SARS-CoV-2 test were compared using the standard laboratory RT-PCR test as a gold standard. Exact Clopper-Pearson 95% confidence intervals were calculated due to estimates being near 1. Kaplan Meier survival analysis was used to compare time to result for the two tests, with log rank testing. Analysis was done using R and STATA version 13.

### Clinical Implementation Study

Following the completion of the COVIDx validation study (May 1^st^ 2020) and demonstration of performance similar to the reference standard test, the hospital switched from standard lab RT-PCR testing to use of SAMBA II for in-hospital testing due to its shorter turnaround time. Twenty SAMBA II machines were operationalised by the CUH POC testing team, each machine capable of performing around 10-15 tests per day. To evaluate the real-world impact of SAMBA on clinical care, we retrospectively gathered data on clinically relevant endpoints from electronic patient records over a ten-day period before and after introduction of the SAMBA test for all patients who underwent COVID-19 testing.

All patients who underwent COVID-19 testing in a 10-day period before and after introduction of the SAMBA II SARS-CoV-2 test were included. Participants were identified from testing reports from the EPIC electronic hospital records system. Clinical and hospital activity data were obtained from the same source.

The primary study outcomes for the implementation study was the median time from admission to definitive bed placement comparing SAMBA assay period with the pre-implementation period. Secondary outcomes were time spent on COVID-19 holding wards, bay closures avoided, proportions of patients in isolation rooms following test, proportions of patients able to be moved to COVID negative open wards following test, and finally whether the test was deemed to have a beneficial impact.

Descriptive analyses of clinical and demographic data are presented as median (IQR) when continuous and frequency (%) when categorical. Difference in continuous variables between the pre and post implementation groups were assessed using the Wilcoxon rank sum tests and difference in categories and proportion were assessed using the Chi-square test or test of proportions. Kaplan Meier survival analysis was used to compare time to definitive bed placement from admission for the two periods, with log rank testing. We fitted Cox proportional hazards models to determine the hazard of placement, after adjustment for age sex, morbidity (defined by a number of scoring systems including quick sequential organ failure assessment score (qSOFA), national early warning score 2 (NEWS2), and Charlson Comorbidity Index (CCI)). In the final multivariable model, estimates of the HRs were determined by including those factors with evidence of an association in the univariable analysis with a *P*-value of < 0.1. Although sex was not significantly associated with time to definitive bed placement in the univariable analysis, it was kept in the final model as it was an a priori specified confounder. Analyses was done using STATA version 13.

### Ethical approval

The protocol was approved by the East of England - Essex Research Ethics Committee. HRA and Health and Care Research Wales (HCRW) approval was received. Verbal informed consent was obtained from all participants or in the case of participants without capacity, from a consultant nominee who was involved in their clinical care but independent from the research team (see supplemental). COVIDx was registered with the ClinicalTrials.gov Identifier NCT04326387. The implementation study was registered as a service evaluation with Cambridge University Hospitals NHS Foundation Trust.

Patients or the public were not involved in the design, or conduct, or reporting, or dissemination plans of our research. There are no plans to directly feedback the results to participants.

## Results

### Validation of SAMBI II SARS-CoV-2 Assay

Of 178 screened patients, 149 met eligibility criteria for inclusion in the clinical trial (Figure 1). Mean age was 62.7 years and 47% were male. 32/149 (21.6%) tested positive by the standard lab RT-PCR test. Mean temperature and respiratory rate were higher in the standard lab RT-PCR positive group (Figure 1). Median duration of symptoms was 3 (IQR 1.75-10.5) and 4 (IQR 2-13) days in standard lab RT-PCR positive and negative participants respectively. There were seven discrepant results between the POC and laboratory assays (7/149) after initial testing (see supplementary methods). Discrepancy analysis concluded that there was one false negative by the POC test, likely related to sampling variation, and no false positives. Cohen’s kappa correlation between the two tests was 0.96, 95% CI (0.91, 1.00). Sensitivity of SAMBA II SARS-CoV-2 test as compared to the standard lab RT-PCR was 96.9% (95% CI 83.8-99.9), with specificity of 99.1% (95.3-99.9), Figure 2. However, since the standard lab RT-PCR had one false negative in a participant with clinical and radiological evidence of disease, the sensitivity and specificity of SAMBA II SARS-CoV-2 test was effectively 97.0% (95% CI 84.2-99.9) and 100% (95% CI 96.9-100) respectively. POC testing was associated with shorter time from sampling to result (Figure 2); median time to result was 2.6 hours (IQR 2.3 to 4.8) for POC testing and 26.4 hours (IQR 21.4 to 31.4) for the standard lab RT-PCR test (p<0.001).

**Figure 1:**
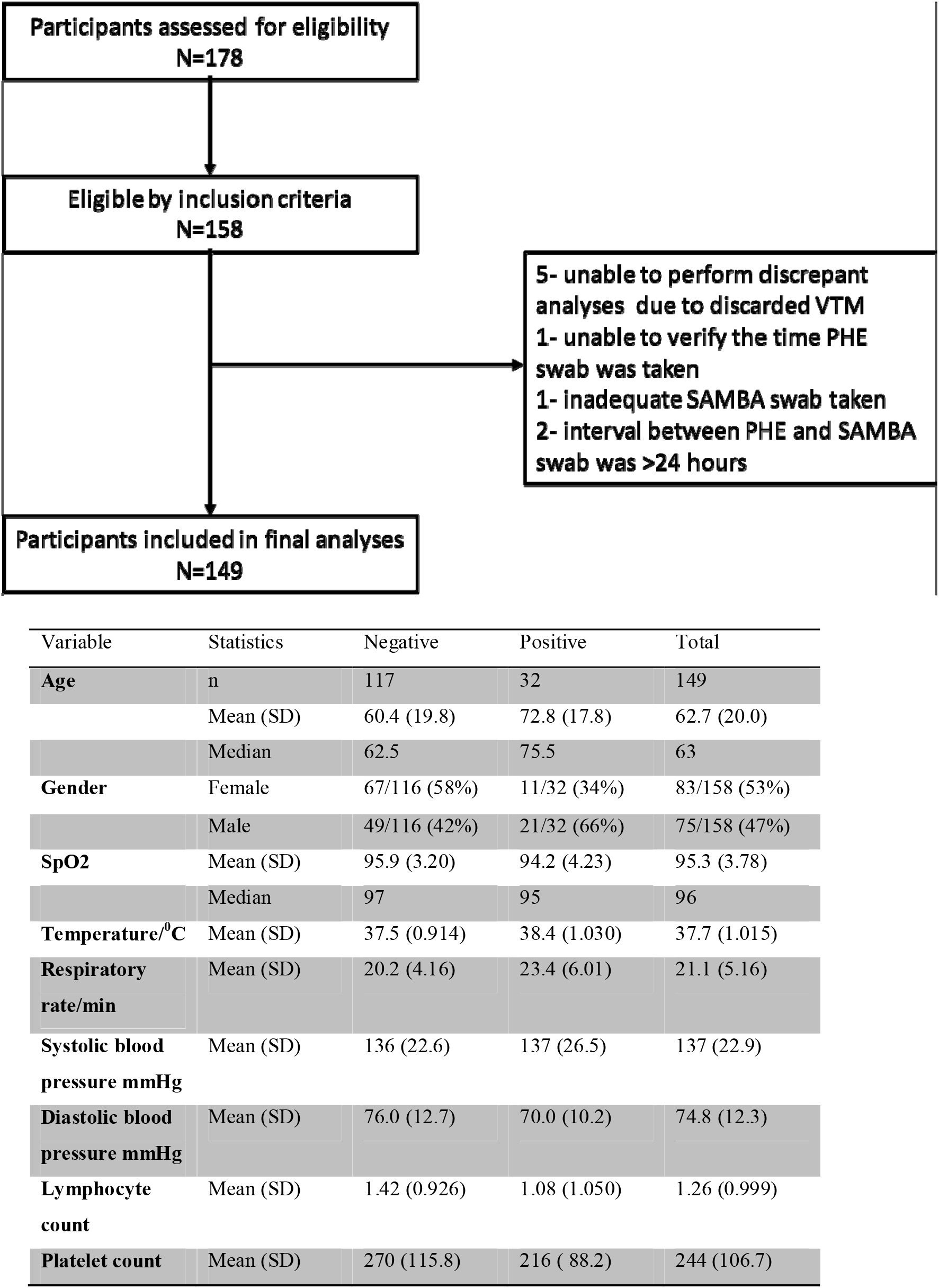
(i) Prospective clinical study flow chart CONSORT diagram. VTM: viral transport medium; PHE: Public Health England. (ii) baseline characteristics of prospective participants in COVIDx trial

**Figure 2.**
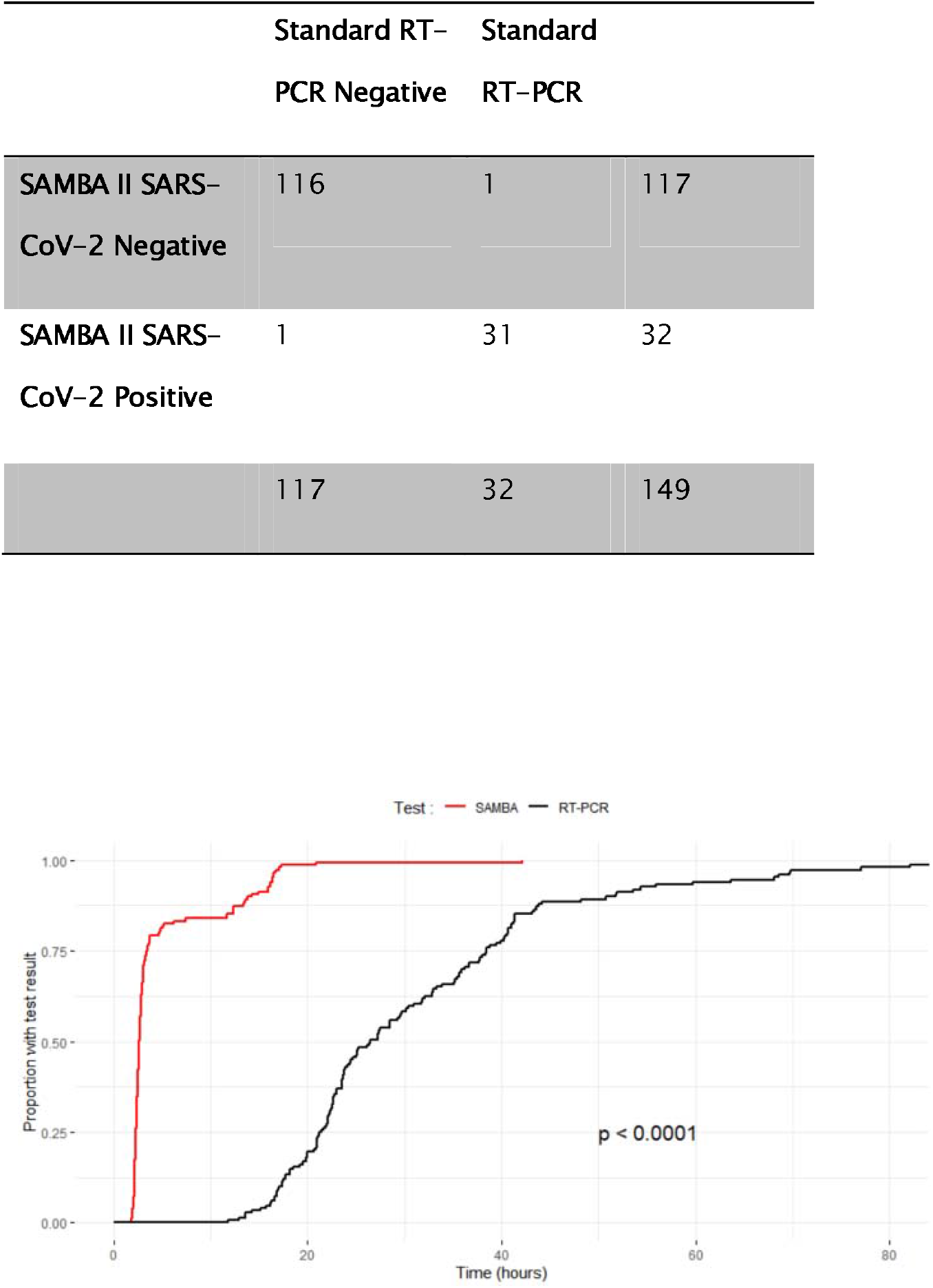
**(top): Accuracy of the SAMBA II SARS-CoV-2 test compared with Standard lab RT-PCR testing; (bottom) Kaplan Meier analysis of time to test result** for Point of Care SAMBA II SARS-CoV-2 and standard RT-PCR test in the COVIDx study. P value shown is for Log rank test.

### SAMBA II SARS-CoV-2 Assay Implementation Study

992 SAMBA II SARS-CoV-2 tests were performed between May 2^nd^ and May 11^th^ inclusive in 913 individuals. The assay was used for the following indications: 59.8% of tests were used for newly hospitalised patients, and the remainder were done for pre-operative screening (11.3%), discharges to nursing homes (10.0%), in-hospital screening of new symptoms (9.7%), screening in asymptomatic patients requiring hospital admission screening (3.8%) and access to interventions such as dialysis and chemotherapy for high risk patients (1.2%) (Table 1). During the implementation phase, median time to result was 3.6 hours (IQR 2.6 to 5.8h). The result from the SAMBA assay was deemed to have a beneficial clinical impact in 77.4% of patients who had the test. (Table 1).

**Table 1:** Clinical and demographic data of 992 tests in 913 patients who had the SAMBA II SARS-CoV-2 test in the post-implementation period. Note that some individuals had multiple admissions each with associated POC tests.

| (N) Individual patients=913/ Tests=992 |  |
| --- | --- |
| Male sex (%) | n=857/913<br>389 (44.6) |
| Median age years (IQR) | n=909/913<br>63 (37-79) |
| Duration of illness days(IQR) | 2 (1-7) |
| SAMBA II SARS-CoV2 result (%) |  |
| Positive | 42 (4.2) |
| Negative | 950 (95.8) |
| Triage at initial assessment (%) | n=966/992 |
| Low risk | 478 (49.5) |
| Medium risk | 387 (40.0) |
| High risk | 101 (10.5) |
| Inpatient Transfer (%) | n=976/992 |
| Yes | 20 (2.0) |
| No | 956 (98.0) |
| Triage following SAMBA II SARS-CoV2 result (%) | n=756/992 |
| Low risk | 600 (79.4) |
| Medium risk | 88 (11.6) |
| High risk | 68 (9.0) |
| Reason for SARS-CoV-2 test | n=970/992 |
| Admission triage and placement | 580 (59.8) |
| In-hospital triage and placement | 94 (9.7) |
| Discharge to nursing home/carers | 97 (10.0) |
| Pre-operative | 110 (11.3) |
| Facilitate other investigations | 12 (1.2) |
| Asymptomatic screening | 37 (3.8) |
| Other | 40 (4.1) |
| Impact of test (%) | N=970/992 |
| Bed placement at admission | 271(28.0) |
| Facilitate discharge to another inpatient facility | 10 (1.0) |
| Release of a side room | 32 (3.3) |
| Expedited discharge | 100 (10.3) |
| Expedited discharge to a nursing home/carers | 58 (6.0) |
| Expedited surgery and other interventions | 128 (13.2) |
| Safe to remain or move to a green ward | 112 (11.6) |
| Avoided a bay closure | 11 (1.1) |
| Facilitated a planned admission | 7 (0.7) |
| No perceived impact | 228 (23.5) |
| Other | 13 (1.3) |

#### Emergency admissions

Rapid SAMBA II SARS-CoV-2 testing was deemed beneficial in 436 (75.8%) tests performed at presentation to ED or the acute admission ward. In the 24.2% where no clinical benefit was derived the reasons for this were: patients being discharged home from ED prior to the result becoming available; patients being triaged and moved to a ward before the results were available; and, in cases of a high clinical index of suspicion of COVID-19, a negative result did not change the initial risk assessment, isolation or clinical management.

#### Pre-operative testing

110 (11.3%) tests were performed prior to surgical procedures, partly for infection control purposes, but mainly to screen patients in light of data demonstrating increased peri-operative mortality associated with COVID-19^24^. POC tests were deemed to have resulted in clinical benefit attributable to the rapid result (Table 3) in 106/110 (96.3%) instances. SAMBA II SARS-CoV-2 testing facilitated surgical interventions including exploratory laparotomy, eye and maxillofacial surgery, solid organ transplants and caesarean sections.

#### Discharge to care home or with a care package

Nursing homes came to be recognised as “hotspots” for COVID-19 transmission and at the end of April 2020 national policy mandated a SARS-CoV-2 swab less than 48 hours before discharge to a nursing home or a setting where an individual was visited by carers. SAMBA II SARS-CoV-2 testing was successfully used to facilitate discharge in 76/96 (79.2%) instances. In the remaining 20.8%, alternative reasons were identified in the discharge pathway that resulted in delays that required another test to meet the hospital’s discharge policy.

#### Prevention of Health Care Associated Infection

94 patients had a SAMBA II POC test carried out for the purpose of in-hospital triage and placement. 81 of these had sufficient data to determine the impact of SAMBA II SARS-CoV-2 test. The test was beneficial in 55.6% (45/81), allowing the patient to remain in a low risk open ward in 68.9% (31/45) instances, movement out of a side room in 17.8% (8/45) and avoiding bay closures in 13.3% (6/45). In the remaining 44.4% (36/81) of instances in which no beneficial impact was found, 7 of these had a previous recent test result of which 2 were known positive, and a SAMBA positive result had no further impact. In 4 instances, the patient had been moved prior to the result returning as clinical suspicion of COVID-19 was high leading to triage prior to the result being known, in 8, there was no documented indication and in the rest SAMBA II SARS-CoV-2 testing did not alter management.

POC testing with negative results allowed a significant increase in the number of patients able to move to ‘green’ non-COVID-19 areas [green status (478/966) 49.5% prior to test and (600/756) 79.4% afterwards, p<0.001]. The numbers in ‘amber’ areas (possible COVID-19) fell reciprocally (Figure 3A) [40% on amber prior to test and 11.6% after, p<0.001], thereby allowing quicker access to potentially life-saving procedures such as CT Angiography or cardiac monitoring (Supplementary material). We observed a concomitant fall in use of single occupancy rooms amongst those tested for new in-hospital COVID-19 symptoms from 30.8% before to 21.2% (p=0.03) after the POC test result (Figure 3B). Eleven bay closures were prevented with POC testing overall, with each bay having an average of six beds.

**Figure 3:**
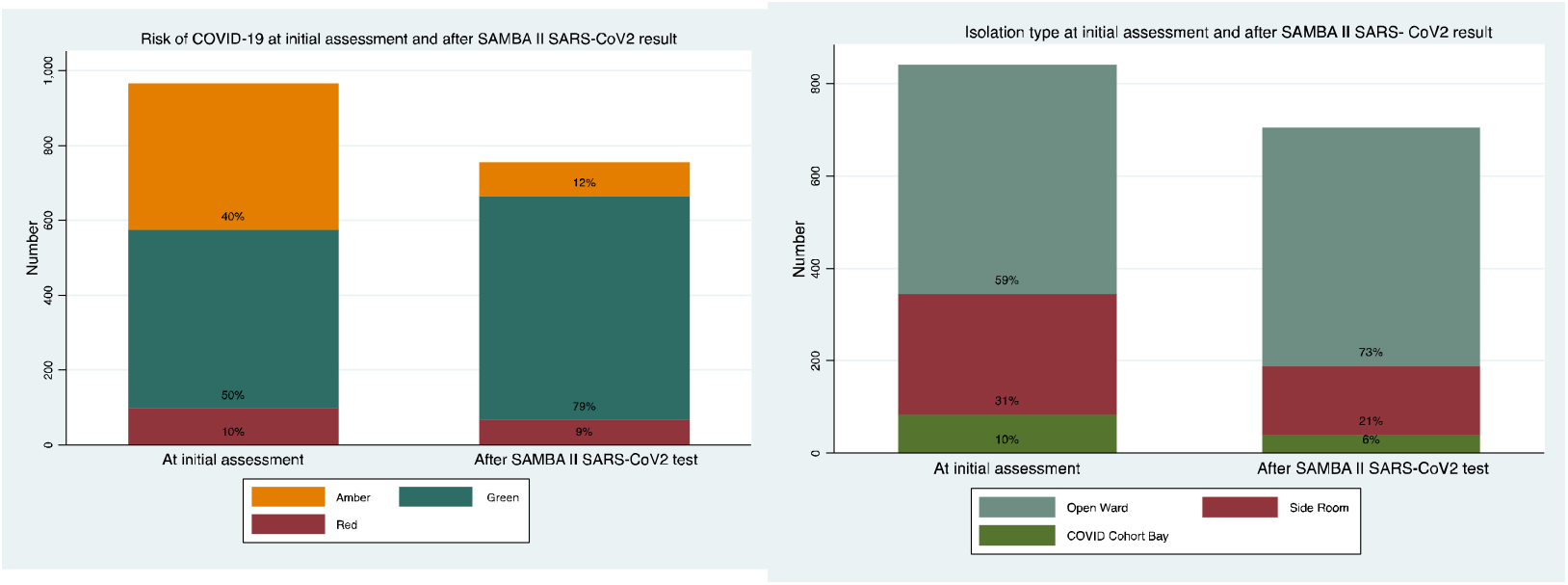
**Impact of SAMBA II SARS-CoV-2 testing on risk stratification** of patients tested with in the post implementation period (left panel, p<0.001 chi squared test) and change in use of single occupancy isolation rooms (right panel, p<0.001 chi squared test). Red, amber and green represent high, medium and low risk clinical areas.

Next we compared clinical outcomes the 10 days prior to and following SAMBA II SARS-CoV-2 introduction. Duplicate tests in the same admission episode were excluded. We identified 561 tests in 388 individuals tested using the standard laboratory RT-PCR in the 10 days prior to SAMBA II SARS-CoV-2 introduction, and compared them with 913 tests done in 799 individuals using the POC test in the 10 days post-SAMBA II SARS-CoV-2 introduction. Demographic characteristics of both groups were similar. Clinical factors were different which reflects the timeline of the pandemic; the proportion of positive tests, mortality and presumed risk of COVID-19 was lower in the post implementation period, (Table 2). Time from sample to test result fell dramatically [35.9 hours (23.8-48.9) to 3.8 hours (2.7-6.0), p<0.0001, Figure 4 shows Kaplan-Meier analysis]. Time to definitive ward move from ED also decreased significantly after SAMBA II SARS-CoV-2 introduction [23.4 hours (8.6-41.9) to 17.1 hours (9.0-28.8), p=0.02, Figure 4 shows Kaplan-Meier analysis]. The Cox proportional hazards regression model showed that even after mutually adjusting for age, sex, quick sequential organ failure assessment score (qSOFA), national early warning score 2 (NEWS2), and Charlson Comorbidity Index (CCI), SAMBA II SARS-CoV-2 test was independently associated with the shorter time to definitive bed placement from admission [HR 1.25 (95% CI 1.02-1.53), p=0.03). Other significant associations were younger age and NEWS2 medium risk score. (Table 3). Finally, mean length of stay on a COVID-19 result wait/holding ward decreased from 58.5 hours to 29.9 hours (p<0.001) compared to the 10 days prior to implementation.

**Figure 4:**
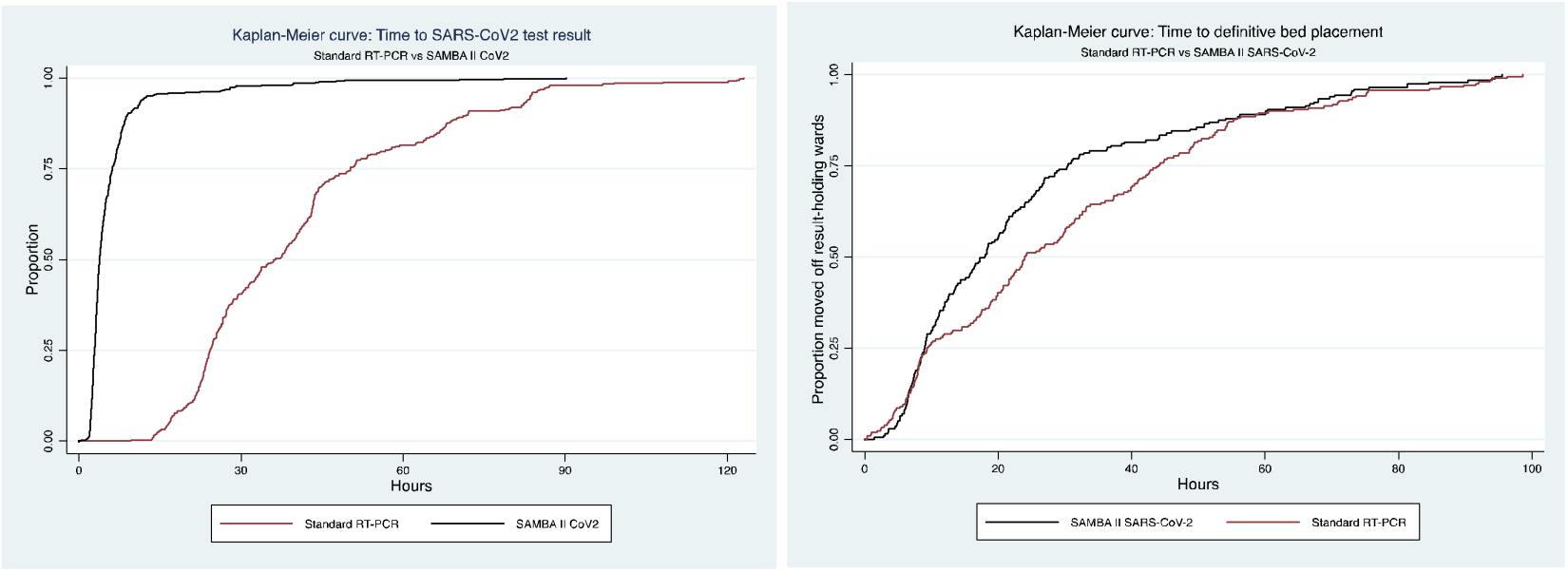
Time to test results (left panel, log rank test p<0.001) and definitive ward move (right panel, log rank test p=0.02) for SAMBA SARS-CoV-2 POC tests in the post implementation period compared to lab RT-PCR in the pre-implementation period.

**Table 2:** **Clinical and demographic data** of patients who had the standard PHE RT-PCR test in the pre-implementation period from 22^nd^ of April 2020 till the 1^st^ of May 2020 and those who had the SAMBA II CoV2 test in the post-implementation period from the 2^nd^ of May 2020 till the 11^th^ of May 2020. Duplicate tests during the same admission period were excluded. qSOFA-Quick sequential organ failure assessment score, NEWS2-National early warning score 2, CCI-Charlson Comorbidity Index. ^a^ Chi-square test ^b^ Wilcoxon rank sum te

|  | Univariable model<sup>†</sup> | Pre-implementation<br>Standard PHE RT-PCR test<br>N= 561 in 388 persons | Post-implementation<br>SAMBA II SARS-CoV-2 test<br>N=913 in 799 persons | Univariable model<sup>†</sup> | P value |
| --- | --- | --- | --- | --- | --- |
| Sex (%) |  |  |  |  |  |
| Male |  | 197 (50.8) | 364 (45.6) |  | 0.10<sup>a</sup> |
| Female |  | 191 (49.2) | 434 (54.4) |  |  |
| Median age years (*IQR*) |  | 63.0 (42.0-79.5) | 61.0 (36.0-78.0) |  | 0.02<sup>b</sup> |
| Acute Admission (%) |  |  |  |  |  |
| Yes |  | 403 (71.8) | 615 (67.4) |  | 0.07<sup>a</sup> |
| No |  | 158 (28.2) | 298 (32.6) |  |  |
| SARS-CoV2 result (%) |  |  |  |  |  |
| Positive |  | 49 (8.7) | 39 (4.3) |  | <0.001<sup>a</sup> |
| Negative |  | 512 (91.3) | 874 (95.7) |  |  |
| Died (%) |  |  |  |  |  |
| Yes |  | 28 (7.2) | 27 (3.4) |  | 0.003<sup>a</sup> |
| Median length of admission days (*IQR*) |  | 4.4 (1.1-10.8) | 2.9 (0.9-7.3) |  | <0.0001<sup>b</sup> |
| Triage at initial assessment (%) |  | N=544/561 | N=856/913 |  |  |
| Low risk |  | 249 (45.8) | 450 (52.6) |  | 0.02<sup>a</sup> |
| Medium risk |  | 244 (44.9) | 349 (40.8) |  |  |
| High risk |  | 51 (9.4) | 57 (6.7) |  |  |
| Median time to test result hours (*IQR*) |  | N=544/561<br>35.9 (23.8-48.6) | N=655/913<br>3.8 (2.7-6.0) |  | <0.0001<sup>b</sup> |
| Median time to definitive bed placement from admission hours (*IQR*) |  | N=160/561<br>23.4 (8.6 to 41.9) | N=267/913<br>17.1 (9.0-28.8) |  | 0.02<sup>b</sup> |
| qSOFA score (%) |  | N=551/561 | N=903/913 |  |  |
| 0-1 |  | 513 (93.1) | 851 (94.2) |  | 0.38<sup>a</sup> |
| 2-3 |  | 38 (6.9) | 52 (5.8) |  |  |
| NEWS2 score (%) |  | N=555/561 | N=906/913 |  |  |
| 0-4 Low risk |  | 407 (73.3) | 711 (78.5) |  | 0.08<sup>a</sup> |
| 5-6 Medium risk |  | 82 (12.9) | 107 (11.8) |  |  |
| >7 High Risk |  | 66 (11.9) | 88 (9.7) |  |  |
| CCI score (%) |  | N=560/561 | N=912/913 |  |  |
| < 4 |  | 470 (83.9) | 782 (85.8) |  | 0.34<sup>a</sup> |
| >=4 |  | 90 (16.1) | 130 (14.2) |  |  |

**Table 3:** **Multivariable analyses** using Cox proportional hazards regression of the effect of SARS-CoV-2 test type on time to definitive bed placement for patients presenting for emergency care in accident and emergency and acute admissions departments. The standard PHE RT-PCR test was used in the pre-implementation period from 22^nd^ of April 2020 till the 1^st^ of May 2020 and the SAMBA II CoV2 test in the post-implementation period from the 2^nd^ of May 2020 till the 11^th^ of May 2020. Only the first test done by each participant in both phases of was included. Only patients who were admitted were included. qSOFA-Quick sequential organ failure assessment score, NEWS2-National early warning score 2, CCI-Charlson Comorbidity Index. ^‡^ Cox regression analyses used except were indicated ^a^ Wilcoxon rank sum test ^b^ Chi-square test § Follow up time in 100 person-hours. 5 Rate per 100 person-hours. * Associations with some evidence against the null.

|  | Events/<br>Follow up<br>time<sup>§</sup> | Rate<sup>δ</sup> | HR (95% CI) | P value | HR (95% CI) | P value |
| --- | --- | --- | --- | --- | --- | --- |
| SARS-CoV-2 Test<br>Standard lab RT-PCR<br>SAMBA SARS-Cov-2 | 211/64<br>201/49 | 3.31 (2.88-3.78)<br>4.04 (3.54-4.67) | 1<br>1.27 (1.05-1.55) | 0.01* | 1<br>1.25 (1.02-1.53) | 0.03* |
| Sex<br>Female<br>Male | 231/63<br>181/50 | 3.64 (3.20-4.15)<br>3.63 (3.14-4.20) | 1<br>0.98 (0.81-1.20) | 0.85 | 1<br>1.01 (0.82-1.23) | 0.94 |
| Age group (years)<br>81-119<br>65-80<br>42-64<br>0-41 | 105/40<br>96/31<br>125/28<br>87/16 | 2.66 (2.19-3.21)<br>3.11 (2.55-3.81)<br>4.54 (3.81-5.41)<br>5.53 (4.48-6.82) | 1<br>1.17 (0.89-1.55)<br>1.84 (1.42-2.39)<br>2.43 (1.82-3.25) | 0.26<br><0.001*<br><0.001* | 1<br>1.29 (0.97-1.71)<br>1.83 (1.40-2.40)<br>2.51 (1.86-2.29) | 0.08<br><0.001*<br><0.001* |
| qSoFA score<br>2-3<br>0-1 | 18/8.2<br>388/100 | 2.20 (1.39-3.50)<br>3.74 (3.39-4.13) | 1<br>1.83 (1.14-2.94) | 0.01* | 1<br>1.54 (0.89-2.66) | 0.12 |
| NEWS2 score<br>>7 High Risk<br>5-6 Medium risk<br>0-4 Low risk | 36/12<br>54/20<br>318/80 | 2.89 (2.08-4.00)<br>2.64 (2.02-3.44)<br>3.98 (3.57-4.44) | 1<br>0.85 (0.55-1.29)<br>1.42 (1.01-2.01) | 0.44<br>0.05* | 1<br>0.58 (0.36- 0.92)<br>0.92 (0.61-1.39) | 0.02*<br>0.69 |
| CCI score<br>≤ 3<br>≥ 4 | 354/94<br>57/19 | 3.76 (3.38-4.17)<br>3.07 (2.36-3.97) | 1<br>0.79 (0.60-1.05) | 0.12 |  |  |

## Discussion

Here we report the an assessment of the validity and impact of rapid molecular SARS-CoV-2 testing for diagnosis of COVID-19 infection in a high-need hospital setting. These data demonstrate that rapid antigen testing can be reliable, accurate and provide clinicians and infection control staff with much quicker results compared to current centralized gold standard assays. Furthermore, we demonstrate that the routine use of this test had a real-world impact on patient triage and hospital bed resource allocation.

The SAMBA II SARS-CoV-2 nucleic acid test was compared to a reference RT-PCR test - the standard of care, using combined nasal/throat swabs from participants attending hospital with a possible diagnosis of COVID-19. Study participants were representative of UK COVID-19 patients^25^, and we found that concordance between the tests was extremely high with Cohen kappa coefficient 0.96. When the standard lab RT-PCR test was referenced as a ‘gold standard’, sensitivity of SAMBA was 96.9% and sensitivity 100%. In the validation stage, median time from swab to result was 2.6 hours for SAMBA II as compared with 26.4 hours for RT-PCR (p<0.001). Importantly, we did identify a single case which was deemed to be a false negative, in comparison to the centralized laboratory assay. However, this patient had both clinical and radiographic findings consistent with COVID-19 disease. Nonetheless, our findings to highlight the importance of COVID-19 triage protocols, which allow for retention of a COVID-19 suspect status, despite a negative nucleic acid test result, in cases with otherwise high pre-test probability of disease^26^.

Patient placement during the COVID-19 pandemic has been a major challenge and has significantly impacted the ability to maintain patient flow and safety in hospital^27^. These data on rapid PCR testing offer one strategy to help address these issues. Our hospital switched from standard lab RT-PCR testing to SAMBA II for in-hospital testing immediately following the end of the validation study, providing an opportunity to prospectively evaluate almost 1000 tests performed over ten consecutive days. Most tests were performed on new admissions to hospital and replicated the large reduction in test turnaround time observed in the clinical validation trial. POC was also used to investigate newly symptomatic patients in hospital to rationalise our limited isolation rooms, and also to rapidly identify new COVID-19 cases with appropriate infection control and prevention of nosocomial outbreaks^11^. Inappropriate isolation is a large drain on staff and resources due to the need for repeated deep cleaning, additional PPE utilisation and the distress and risk to patients from repeated bed moves^28^. When we performed implementation impact analysis using ten day windows either side of hospital-wide assay implementation, we found that time to definitive ward move from ED decreased significantly after SAMBA II SARS-CoV-2 introduction, and length of stay on the main holding ward where test results were awaited also fell significantly, consistent with more rapid and accurate patient movement. Similarly, we observed a significant increase in the availability of isolation and single occupancy rooms following POC introduction, and also patients testing negative were able to be placed in low risk areas of the hospital and have interventions/procedures expedited. Finally, we found that 11 ward closures were prevented in the ten-day post implementation phase by having negative tests in symptomatic hospital patients. Closed surgical bays in particular can result in cancellations of operations, as well as significant financial losses to hospitals. Following this analysis, hospital guidelines will be adapted to recommend waiting for SAMBA test results before moving patients into isolation or closing bays.

SAMBA II SARS-CoV-2 test is being implemented in a very limited number of hospitals, but there is urgent need for similar capacity in care homes, prisons and possibly other establishments. A rapid POC platform also has the potential to reduce disparities between secondary and tertiary medical centres that have specialised virology laboratories, and ensure equitable access to timely SARS-CoV-2 testing results. SAMBA II machines are already in use in Uganda, Zimbabwe and Kenya for HIV testing and monitoring. If scale up can be achieved in those settings, rapid testing could be vital for controlling COVID-19 in sub-Saharan Africa^8^ and our data will inform their optimal use^29^.

### Limitations

The assay validation component was limited by the fact that the same swab could not be tested on the two platforms being compared. This raised an issue of two separate samples being tested on the two assays. Nonetheless, we identified only 2 cases where the sampling explained discrepant results. In addition, the SAMBA II SARS-CoV-2 test is not able to give viral load or cycle threshold values for more nuanced analysis. Results of the validation can be generalized to hospitalized suspects of COVID-19 with symptom of disease, but we did not assay validity in asymptomatic or outpatients with mild symptoms. Similarly, our results included dual swabs of the oral and naso-pharynx and should be interpreted with those methods mind.

The implementation study was a non-randomized, controlled pre-post intervention design, and thus the effects seen cannot be fully causally attribute to the implementation of the assay. However, our findings are plausible, consistent across multiple measures, and process measures (e.g. turnaround time) are supportive of the more downstream measures assessed. Moreover, for our primary outcome, we conducted multivariable adjustment including clinical and demographic indicators, which demonstrated a persistent benefit in hazard of time to emergency room discharge. Moreoever, the implementation phase took place six weeks into the UK lockdown, at a time when the rate of new infections had reduced substantially across the country. Nonetheless, the study highlights the importance of rapid test results in the COVID-19 era, regardless of the outcome of the test results.

Finally, our study did not estimate costs of the cost effectiveness of the implementation strategy. The utilisation of rapid assays in acute settings for other respiratory viruses has been shown to be cost effective^30^. Given the morbidity and mortality associated with COVID-19, as well as the disruption that this pandemic has placed on healthcare provision, we anticipate future assessments of the cost implications of SAMBA II SARS-CoV-2 implementation in regards to delayed discharge, nosocomial transmission and unnecessary use of personal protective equipment

In summary, our data suggest that implementation of rapid testing for SARS-CoV-2 could have a critical impact on hospital management of suspected COVID-19 cases. Future studies should assess the long-term implications, resources, and clinical efficiency of rapid assay implementation, particularly in the context of influenza and norovirus seasons.

## Data Availability

data can be obtained from the corresponding author by email at

## Acknowledgements

We would like to thank the staff and patients at CUH NHS Foundation Trust.

## Data sharing

raw anonymised data are available from the corresponding author

## The CITIID-NIHR COVID BioResource Collaboration

### Principal Investigators

Stephen Baker, John Bradley, Gordon Dougan, Ian Goodfellow, Ravi Gupta, Paul J. Lehner, Paul Lyons, Nicholas J. Matheson, Kenneth G.C. Smith, Mark Toshner, Michael P. Weekes

### Clinical Microbiology & Public Health Laboratory (PHE)

Nick Brown, Martin Curran, Surendra Palmar, Hongyi Zhang, David Enoch.

### Institute of Metabolic Science, University of Cambridge

Daniel Chapman

### Cambridge University Hospitals NHS Foundation Trust, Cambridge, UK

Ashley Shaw

### NIHR Cambridge Clinical Research Facility

Vivien Mendoza, Sherly Jose, Areti Bermperi, Julie Ann Zerrudo, Evgenia Kourampa, Caroline Saunders, Ranalie de Jesus, Jason Domingo, Ciro Pasquale, Bensi Vergese, Phoebe Vargas, Marivic Fabiculana, Marlyn Perales, Richard Skells.

### Cambridge Cancer Trial Centre

Lee Mynott, Elizabeth Blake, Amy Bates, Anne-laure Vallier, Alexandra Williams, Richard Skells, David Phillips, Edmund Chiu, Alex Overhill, Nicola Ramenante, Jamal Sipple, Steven Frost, Helena Knock, Richard Hardy, Emily Foster, Fiona Davidson, Viona Rundell, Purity Bundi, Richmond Abeseabe, Sarah Clark, Isabel Vicente

